# Cutaneous *Leishmania mexicana* Infections in the United States: Defining Strains Through Endemic Human Pediatric Cases in Northern Texas

**DOI:** 10.1101/2024.01.11.23300611

**Authors:** Binita Nepal, Clare McCormick-Baw, Karisma Patel, Sarah Firmani, Dawn M. Wetzel

## Abstract

Over a six-month span, three patients under five years old with cutaneous leishmaniasis presented to the Pediatric Infectious Diseases Clinic at the University of Texas Southwestern Medical Center/Children’s Health Dallas. None had traveled outside of the United States (US); all had confirmed *L. mexicana* infections by PCR. We provide case descriptions and images to increase the awareness of this disease among US physicians and scientists. Two patients responded to fluconazole, but one required topical paromomycin. Combining these cases with guidelines and our literature review, we suggest that: 1) higher doses (ten-twelve mg/kg/day) of fluconazole should be considered in young children to maximize likelihood and rapidity of response and 2) patients should transition to alternate agents if they do not respond to high-dose fluconazole within six weeks. Furthermore, and of particular interest to the broad microbiology community, we used samples from these cases as a proof-of-concept to propose a mechanism to strain-type US-endemic *L. mexicana.* For our analysis, we sequenced three housekeeping genes and the internal transcribed sequence 2 of the ribosomal RNA gene. We identified genetic changes that not only allow us to distinguish US-based *L. mexicana* strains from strains found in other areas of the Americas, but also establish polymorphisms that differ between US isolates. These techniques will allow documentation of genetic changes in this parasite as its range expands. Hence, our cases of cutaneous leishmaniasis provide significant evolutionary, treatment and public health implications as climate change increases exposure to formerly tropical diseases in previously non-endemic areas.

**IMPORTANCE:** Leishmaniasis is a parasitic disease that typically affects tropical regions worldwide. However, the vector that carries *Leishmania* is spreading northward into the United States (US). Within a six-month period, three young cutaneous leishmaniasis patients were seen at the Pediatric Infectious Diseases Clinic at The University of Texas Southwestern/Children’s Health Dallas. None had traveled outside of north Texas. We document their presentations, treatments, and outcomes and compare their management to clinical practice guidelines for leishmaniasis. We also analyzed the sequences of three critical genes in *Leishmania mexicana* isolated from these patients. We found changes that not only distinguish US-based strains from strains found elsewhere, but also differ between US isolates. Monitoring these sequences will allow tracking of genetic changes in parasites over time. Our findings have significant US public health implications as people are increasingly likely to be exposed to what were once tropical diseases.

## INTRODUCTION

Leishmaniasis is caused by obligate intracellular parasites of the genus *Leishmania*, which are spread by *Lutzomyia* and *Phlebotomus* sand flies. A spectrum of human illness results, including cutaneous, mucocutaneous, and visceral disease.

Disease manifestations are governed by the infecting parasite species and host immune status (*1*).

Generations of medical providers and infectious diseases specialists have been taught that leishmaniasis occurs in tropical and subtropical regions, and not the United States (US). Such viewpoints are reflected in published distributions of leishmaniasis (*2*). However, over time, the sand fly vector’s distribution has expanded into previously non-endemic areas (*3*), and leishmaniasis is well-documented in US animals (*4*). In addition, multiple articles have described human acquisition of cutaneous leishmaniasis within Oklahoma and Texas, where leishmaniasis is now a reportable disease (*5*). In fact, a cross-sectional observational study indicated that 41/69 cases (59%) of cutaneous leishmaniasis diagnosed from 2007-2017 in Texas were acquired locally rather than from travel (*6*). Nevertheless, it is common for US providers to contemplate a leishmaniasis diagnosis in only patients with a history of foreign birth, travel, or military service, which in turn often excludes pediatric patients from consideration.

Within a 6-month period, 3 patients with cutaneous leishmaniasis presented to the Pediatric Infectious Diseases (ID) Clinic at the University of Texas Southwestern Medical Center (UTSW)/Children’s Health Dallas. These cases reflect this disease’s changing epidemiology, since clinical isolates from these patients had genetic polymorphisms that have been documented in Texan strains of *L. mexicana*. Furthermore, we sequenced three *Leishmania* housekeeping genes and identified significant differences between our clinical isolates, providing us with a novel mechanism to type *L. mexicana* strains. Monitoring these sequences could permit documentation of genetic variation among *Leishmania* isolates as this parasite expands its geographic range in the US.

## RESULTS

### Patient presentations

3 cases of cutaneous leishmaniasis acquired in Texas were referred to the Pediatric ID Clinic at UTSW/Children’s Health Dallas over a six month period by their treating dermatologists. Interestingly, none of these patients’ family members had lesions. Descriptions of these cases follow (**Figure 1**).

**Figure 1:**
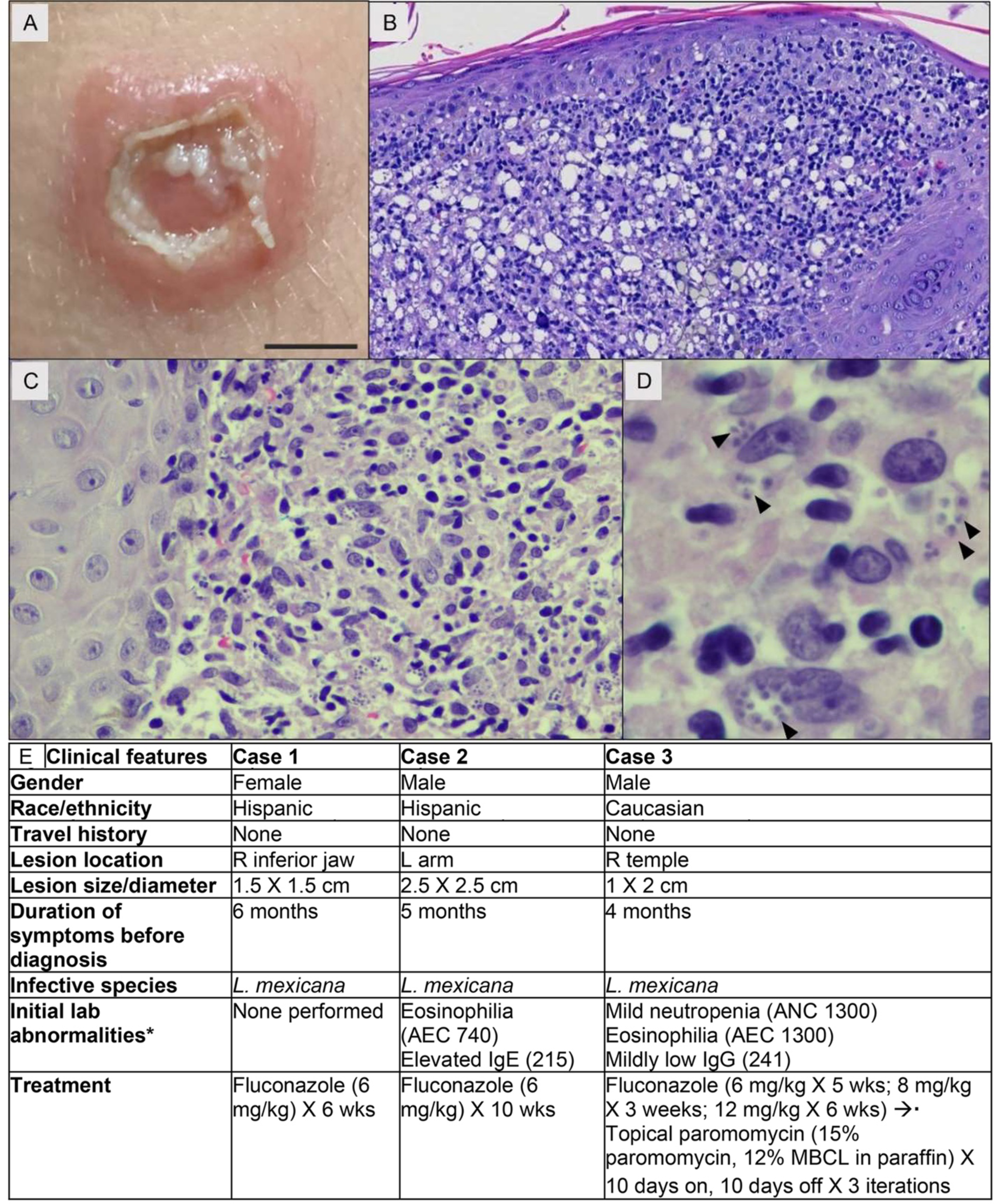
Clinical features of patients presented. **A-D)** Clinical and histopathological evaluation of patient 2. **A)** Approximately 2.5 cm lesion on the left arm clinically consistent with cutaneous leishmaniasis. Scale bar = 1 cm. **B)** Granulomatous inflammation with numerous histiocytes (100x magnification, H&E). Sample shared from Sagis Diagnostics, Houston, TX. **C)** Small, round amastigotes are visible within histiocytes in the reticular dermis (500x magnification, H&E). **D)** Amastigotes within histiocytes with visible kinetoplasts at arrowheads (1000x magnification, oil immersion, H&E). **E)** Patient demographics, signs and symptoms, and laboratory findings. *Normal studies in Case 2 included CBC, remainder of differential, C-reactive protein, and chemistries including liver function panel. Normal studies in Case 3 included CBC, remainder of differential, C-reactive protein, chemistries including liver function panel, IgA, IgM, and IgE. All laboratory abnormalities resolved after treatment. ANC, Absolute neutrophil count. AEC, Absolute eosinophil count.

#### Case 1

A 0-5 year old girl reported a non-healing nodular lesion on her right jawline that had been present for months. She had not traveled outside north Texas. The lesion had not responded to antibiotic therapy and worsened with steroids. A biopsy showed a granulomatous infiltrate within the dermis, with amastigotes observed inside histiocytes, diagnostic of cutaneous leishmaniasis. Genetic analysis performed at the Centers for Disease Control and Prevention (CDC) demonstrated *L. mexicana* infection. She was started on 6 mg/kg/day of fluconazole and, with 6 weeks of treatment, the lesion essentially resolved.

#### Case 2

A few months later, a 0-5 year-old boy who had no travel history developed a non-healing nodular lesion on his left arm. It did not respond to antibiotics and ulcerated after steroids were administered. He was seen in the ID clinic and had a nearly 2.5 cm-diameter ulcer (**Fig 1A**). Biopsy demonstrated granulomatous inflammation with innumerable amastigotes within histiocytes (**Fig 1B-D**). PCR amplification of parasite DNA performed through the CDC detected *L. mexicana*. He received 6 mg/kg/day of fluconazole and required 10 weeks of therapy for lesion resolution.

#### Case 3

A 0-5 year old boy developed a nodular facial lesion that did not improve with antibiotics and worsened with steroids. He had never traveled outside north Texas/southern Oklahoma; however, he had been taken for daily walks at sunset at a lake near his home. A biopsy and genetic sequencing performed at the CDC demonstrated *L. mexicana* infection, leading to ID referral approximately 1 month after patient 2. He was treated with fluconazole at 6 mg/kg/day, which was increased to 12 mg/kg/day after minimal clinical response. When his lesion failed to improve on fluconazole, he was given topical paromomycin, compounded as in **Figure 1E** (*7*). After several months, his lesion resolved.

### Genetic sequencing of patient isolates

Since there appeared to be an elevated incidence of local *Leishmania* infections, we explored whether there was genetic variability in parasites from clinical samples. With permission from patients 2 and 3, we obtained DNA from lesions for molecular testing (termed Tx2 and Tx3, **Figure 2** and **Supplemental Material**). Unfortunately, a sample from the initial patient (patient 1) was not available for further study. We then proceeded with molecular testing as follows.

**Figure 2.**
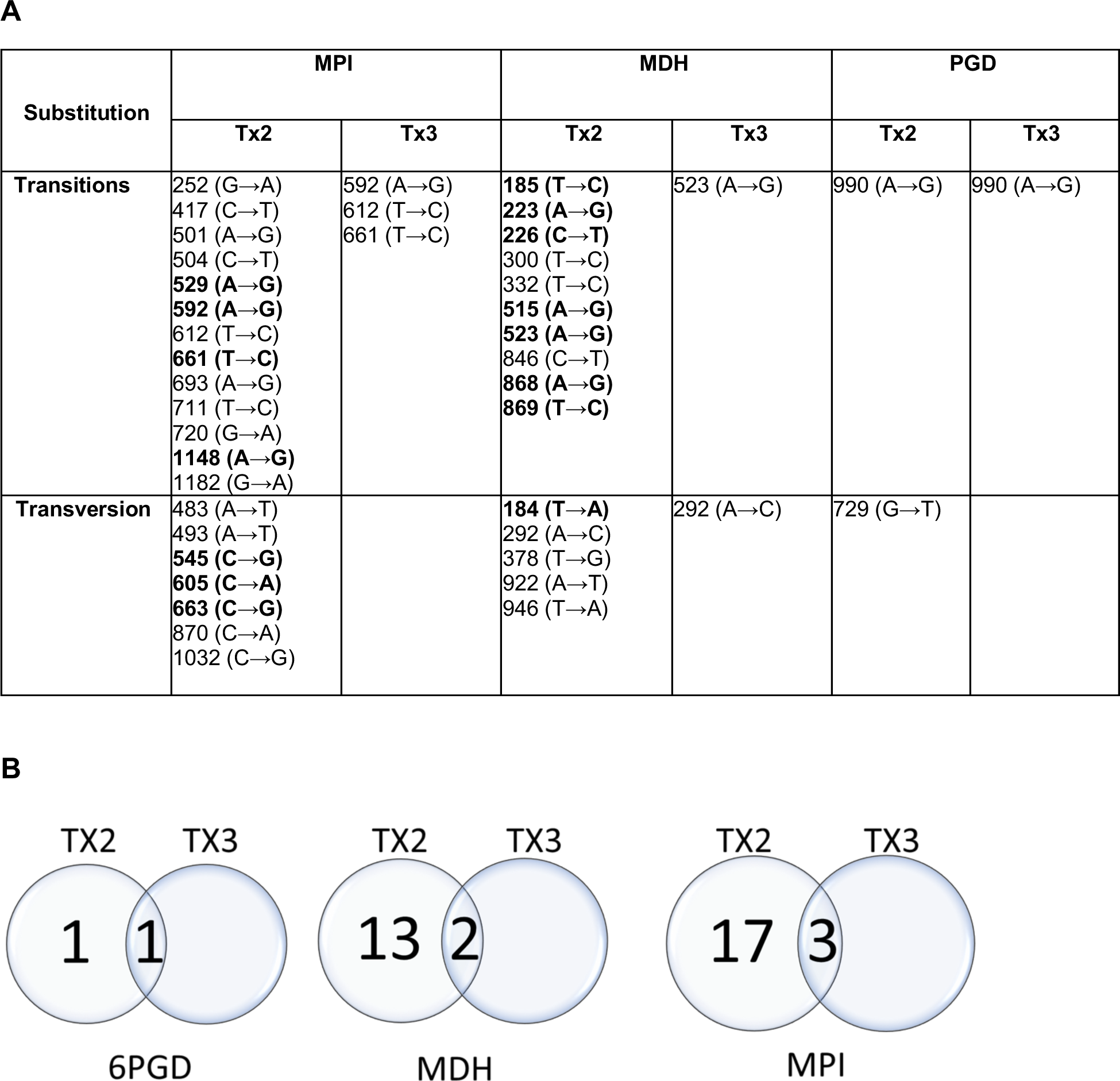
Genetic analysis of clinical isolates. **A**) SNPs reported after MLSA analysis. The nonsynonymous SNPs are in bold. **B)** Venn diagram showing SNPs reported on genes analyzed by MLSA.

#### ITS2 sequencing

The CDC sequences the ribosomal RNA-internal transcribed spacer 2 (ITS2) region to identify the infecting *Leishmania* species, since having this information is needed for appropriate therapeutic decisions (*7*). Texas-specific ITS2 polymorphisms in *L. mexicana* have been described, specifically A è C647 and T è C649 (*8*). Our sequence analysis confirmed that these ITS2 polymorphisms were seen in both of our isolates (**Supplemental Figure and Table**).

#### Multilocus Sequence Analysis (MLSA)

We next explored whether there was intraspecies variation among isolates using multilocus sequence analysis (MLSA). We selected a panel of 3 housekeeping genes: mannose phosphate isomerase (MPI), malate dehydrogenase (MDH) and 6 phosphogluconate dehydrogenase (6PGD). We used gene-specific primers for amplification and Sanger sequencing to identify synonymous/nonsynonymous SNPs in coding regions. Of note, our primers did not amplify genetic material from uninfected human samples.

We found that 6PGD was the most conserved gene, with the least number of SNPs in both isolates. Interestingly, the MDH and MPI genes of Tx2 had significantly diverged from reference strains. In fact, the MDH enzyme in Tx2 was prematurely truncated (by 9 aa at the C terminus) due to transversion 922 (A→T) (**Figure 2, Supplemental Table**). Tx3 also had several SNPs in the MDH and MPI genes that did not match reference strains (**Figure 2, Supplemental Table**). Hence, MLSA analysis of these three metabolic genes may provide a unique molecular epidemiological mechanism to study genetic divergence in *L. mexicana*.

## DISCUSSION

Here, we have documented 3 cases of cutaneous leishmaniasis due to *L. mexicana* acquired by pediatric patients within northern Texas. Since our described patients had normal immune systems and did not have infected family members, we believe they were individually exposed to sandflies outside their homes. Despite their relatively small number, our cases have significant bearing on US public health initiatives, as the likelihood of US patient exposure to what were formerly geographically-constrained tropical diseases will likely continue to increase as shifting vector distribution and other drivers continue (*3*). In addition, we have provided a simple means to type strains of *L. mexicana* endemic to the United States. We propose that our methodology can be used to monitor genetic changes in *Leishmania* as the range of this parasite expands.

Our cases provide multiple therapeutic imputations for treating physicians in the US. Per Infectious Disease Society of America (IDSA) guidelines, treating single lesions from non-disseminating cutaneous species is optional, even in pediatric patients (*7*). However, we treated our patients for cosmetic reasons, since each had facial or large lesions. The referring dermatologists were hesitant to utilize local cryotherapy or brachytherapy (standard for single *L. mexicana*-induced lesions) due to patient age. IDSA guidelines indicate that employing fluconazole is reasonable for US pediatric cutaneous leishmaniasis patients (*7*). Therefore, our patients were initially treated with oral fluconazole, which has a ∼50% success rate among all species (*9, 10*); 2 of our patients responded. Fluconazole is generally well-tolerated, has good oral bioavailability, and achieves epidermis/dermis concentrations above plasma levels (*11, 12*). A dose of 6 mg/kg typically is recommended for pediatric cutaneous leishmaniasis (*7*). However, for serious fungal infections, many authors propose higher doses (12 mg/kg) in children based on fluconazole’s lower half-life, higher volume of distribution, more rapid clearance, and minimal side effect profile in pediatric patients, to prevent treatment failure or relapse (*13*), (*14*). Combining this literature with our experience, initiating higher doses (10-12 mg/kg/day (*15*)) in younger (≤ 5-year-old) pediatric leishmaniasis patients may be worthwhile to maximize likelihood and rapidity of response. Furthermore, we note our third patient did not respond to months of fluconazole therapy. Consistent with IDSA guidelines, we propose that pediatric leishmaniasis patients be transitioned to alternate agents if they do not begin to respond to high-dose fluconazole in 4-6 weeks (*7*). Notably, the topical paromomycin employed in patient 3 can be difficult to acquire in the US, as it requires compounding by a willing pharmacy and investigational drug applications, so we did not use it initially in our patients.

We also determined that our patients’ *Leishmania* isolates have undergone significant genetic changes that may offer insights into evolutionary biology. Notably, it is now more common for patients in Texas to acquire leishmaniasis locally than from travel (*5, 6*). The CDC has described a specific polymorphism in the ITS2 sequence of *L. mexicana* in Texas-endemic strains; we replicated these results in our patients’ isolates. Furthermore, we provide a mechanism for strain-typing isolates of this newly-endemic parasite using 3 metabolic genes. Our results demonstrate the power of MLSA analysis as a simple yet important molecular epidemiological tool to study *Leishmania* genetic divergence within the US. Since clusters of leishmaniasis cases have been described, our technique could potentially be used to characterize outbreaks. The large number of genetic changes that we found using a limited number of isolates suggests that Texas-specific strains of *L. mexicana* may be under significant evolutionary selection pressure, which in turn may lead to novel mutations. We plan to continue to monitor available isolates for this possibility over time.

Although cases of leishmaniasis certainly have been described in the United States, our case discussions have typically been met with surprise from clinicians and scientists alike. One factor that may lead to this underappreciation of endemicity is that leishmaniasis is a reportable disease in Texas, but not in other states (*6*). We would argue that leishmaniasis should be a reportable disease nationwide so that its incidence and distribution can be monitored by public health officials. Furthermore, we propose that such information could be collected in conjunction with our strain-typing techniques to characterize ongoing genetic variation in this parasite as its endemic area expands. Hence, the cases of cutaneous leishmaniasis described here have significant microbiological, treatment and public health implications.

## METHODS

### Molecular analysis/Strain typing

For patient 2 in this report, a clinical sample (termed Tx2) was obtained and briefly cultured at UTSW as promastigotes in Schneider’s Insect cell media (Sigma, S9895) supplemented with 20% FBS (GeminiBio, 100-106) and 1% penicillin-streptomycin (Sigma, P4333). Time in culture was limited to minimize genetic changes *in vitro*. DNA was extracted using the DNAeasy Blood and tissue Kit (Qiagen, 69504) according to manufacturer’s instructions. For patient 3 (Tx3), we received extracted DNA from the CDC (Thanks to Yvonne Qvarnstrom, CDC), which had been generated in a similar manner to Tx2. Extracted DNA was used to amplify the rRNA-internal transcribed spacer (ITS2) region for species identification (**Supplemental Figure**) and three housekeeping genes: mannose phosphate isomerase (MPI), malate dehydrogenase (MDH) and 6 phosphogluconate dehydrogenase (6PGD) with gene specific primers for *L. mexicana*. Sequences for all primers are in the **Supplemental Table**. The CDS region for these genes was then used for MLSA to explore genetic variation among isolates. Briefly, each gene was amplified with high fidelity DNA polymerase Primestar Max (Takara Bio, RO45A) using forward and reverse primers binding in upstream and downstream coding regions, respectively. PCR amplified fragments were Sanger sequenced in the UTSW McDermott Center Sequencing Core facility using Life Technologies® Dye Terminator 3.1 chemistry and 3730XL Genetic Analyzers.

### Single nucleotide polymorphism (SNP) analysis

Each gene was sequenced bidirectionally using 2 sets of forward (ExtF, IntF) and reverse (ExtR and IntR) primers. ExtF and ExtR bind outside of the coding region and IntF and IntR bind in the coding region. Due to the sequence variability found in the Tx2 MPI gene, multiple internal primers were used. SNP analysis was performed using the sequence alignment tools EMBOSS needle and Clustal Omega (www.ebi.ac.uk).

## Supporting information

Supplemental Material aligned genetic sequences

## Data Availability

All data produced in the present study are contained in the manuscript or will be available upon reasonable request to the authors.

## ACKNOWLEDGEMENTS

We are grateful to staff members in the UTSW/Children’s Health Dallas Pediatric Infectious Diseases clinic for their flexibility in patient scheduling to ensure that these leishmaniasis patients saw Dr. Wetzel. We thank the Texas Department of Health for discussing these cases with us when we reported them, members of the UT Southwestern and global parasitology communities for their support and interest in this experience, and the Pediatric ID Division at UT Southwestern for providing helpful advice. We also thank Dr. Yvonne Qvarnstrom at the CDC for assisting us with acquiring leishmanial DNA from patient 3. Dr. Nepal is funded by American Heart Association Postdoctoral Fellowship 916875. Dr. Wetzel was supported by a Children’s Clinical Research Advisory Committee (CCRAC) Early Investigator Award, a 2019 Harrington Scholar-Innovator Award, NIH R01 AI146349, a Welch Grant for Chemistry (I-2086) and funds from the UTSW Department of Pediatrics. The funders did not play a role in the writing of this manuscript. Reporting of this small case series (≤ 3 patients) was exempt from UTSW/Children’s Health Dallas-IRB approval. However, the patients in this study provided permission for images to be used and for clinical samples to be taken; these activities were approved by the IRBs at both UTSW and Children’s Health (STU-2019-0754).

## SUPPLEMENTAL MATERIAL

### Supplemental Methods, Results and Discussion

Additional specific details regarding genetic sequencing of the clinical isolates from these patients are described below. Of interest, this analysis can be done after sampling lesions with a flocked nylon swab, which provides a less invasive means of diagnosis in children. All sequences described in this manuscript have been submitted to TriTryp DB (<u>TriTrypDB)</u>.

#### ITS2

We performed sequence analysis on the rRNA-internal transcribed spacer 2 (ITS2) region, which shows a previously reported Texas-specific polymorphism, C647 and C649, on both Tx2 and Tx3. For this analysis, FJ948434 was used as a reference strain. However, alignments of the FJ948434 ITS2 sequence with 25 publicly available non-US *L. mexicana* ITS2 sequences were identical. Isolates’ ITS2 sequences are shown in the **Supplemental Figure**, and primer sequences are contained in the **Supplemental Table**.

#### Metabolic enzymes

For the three metabolic enzymes sequenced, *Leishmania mexicana* MHOM/GT/2001/U1103 (Guatemala) was used as a reference strain, since it is the *L. mexicana* strain with the most widely available genomic sequence (*1*). *MDH:* SNP analysis in the CDS of the MDH gene showed a total of 15 SNPs in Tx2. We identified nine base transitions: 185 (T→C), 223 (A→G), 226 (C→T), 300 (T→C), 332 (T→C), 515 (A→G), 523 (A→G), 846 (C→T), 868 (A→G), 869 (T→C), and five base transversions, 184 (T→A), 292 (A→C), 378 (T→G), 922 (A→T), 946 (T→A). Tx3 had two SNPs, a base transition at 523 (A→G) and a transversion at 292 (A→C). We note that SNPs 184 (T→A), 185 (T→C), 223 (A→G), 226 (C→T), 515 (A→G), 523 (A→G), 868 (A→G), 869 (T→C) were nonsynonymous and resulted in amino acid changes (**Figure 2, Supplemental Table**). Of particular interest, MDH in Tx2 was prematurely truncated (by 9 aa at the C terminus) due to transversion 922 (A→T). However, this truncation would not be expected to affect its enzymatic function (*2*).

*PGD:* Similarly, we identified 2 SNPs on coding region of PGD gene of Tx2, transversion at 729 (G→T) and single transition 990 (A→G). Tx3 had a single base transition at 990 (A→G).

*MPI:* The MPI gene for the Tx3 isolate had 3 base transitions, 592 (A→G), 612 (T→C), 661 (T→C). On the other hand, for Tx2, the MPI gene had 20 SNPs: transitions 252 (G→A), 417 (C→T), 501 (A→G), 504 (C→T), 529 (A→G), 592 (A→G), 612 (T→C), 661 (T→C), 693 (A→G), 711 (T→C), 720 (G→A), 1148 (A→G), 1182 (G→A) and transversions: 483 (A→T), 493 (A→T), 545 (C→G), 605 (C→A), 663 (C→ G), 870 (C→A), 1032 (C→G). SNPs 529 (A→G), 545 (C→G), 592 (A→G), 605 (C→A), 661 (T→C), 663 (C→G), 1148 (A→G) were nonsynonymous.

## Supplemental Figure

**Supplemental Figure.**
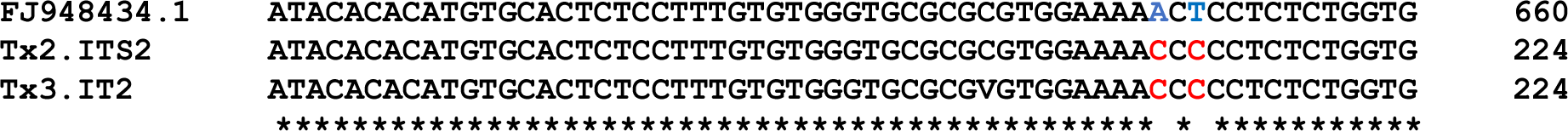
Sequences of the relevant ITS2 region in a representative non-US reference strain (FJ948434.1), compared to Tx2 and Tx3.

## Supplemental Table

**Supplemental Table.**
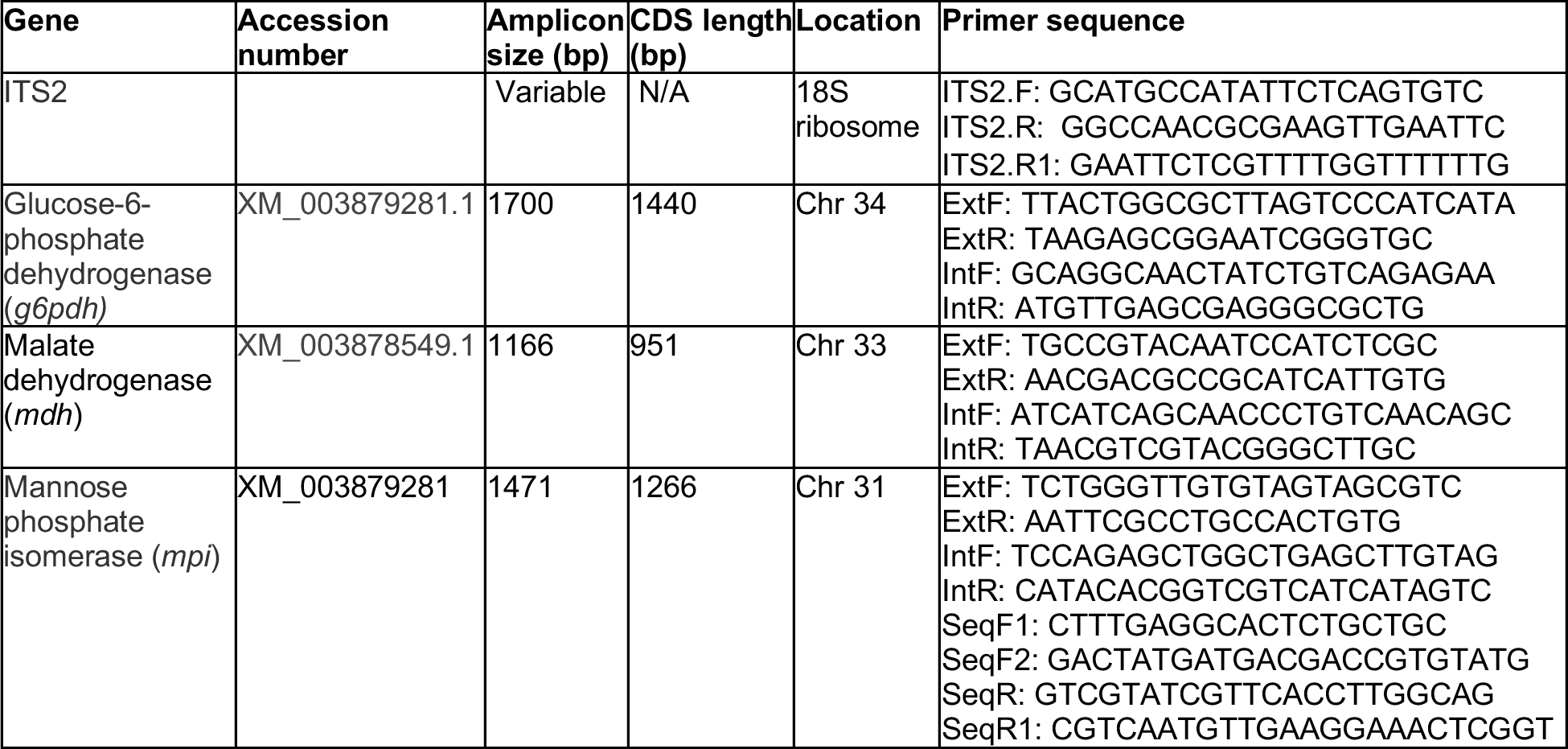
Genes and primer sequences used for MLSA analysis.

