## Supplemental Material aligned genetic sequences for "Cutaneous *Leishmania mexicana* Infections in the United States: Defining Strains Through Endemic Human Pediatric Cases in Northern Texas"

**MDH** CLUSTAL O(1.2.4) multiple sequence alignment

```
Ref.MDH      ATGCGCCGCTCTCAGGCATCCTTCTTCCGTGTCGCTGTGCTCGGTGCTGCCGGTGGCATC   60
Tx2.MDH      ATGCGCCGCTCTCAGGCATCCTTCTTCCGTGTCGCTGTGCTCGGTGCTGCCGGTGGCATC   60
Tx3.MDH      ATGCGCCGCTCTCAGGCATCCTTCTTCCGTGTCGCTGTGCTCGGTGCTGCCGGTGGCATC   60
             ************************************************************

Ref.MDH      GGCCAGCCCTTGTCGCTCCTTCTCAAAAATAACAAGTACGTGAAGGAGCTGAAACTGTAC   120
Tx2.MDH      GGCCAGCCCTTGTCGCTCCTTCTCAAAAATAACAAGTACGTGAAGGAGCTGAAACTGTAC   120
Tx3.MDH      GGCCAGCCCTTGTCGCTCCTTCTCAAAAATAACAAGTACGTGAAGGAGCTGAAACTGTAC   120
             ************************************************************

Ref.MDH      GATATCAAGGGCGCCCCTGGTGTGGCTGCAGACCTCTCCCACATCTACACCCCAGCGAAG   180
Tx2.MDH      GATATCAAGGGCGCCCCTGGTGTGGCTGCAGACCTCTCCCACATCTACACCCCAGCGAAG   180
Tx3.MDH      GATATCAAGGGCGCCCCTGGTGTGGCTGCAGACCTCTCCCACATCTACACCCCAGCGAAG   180
             ************************************************************

Ref.MDH      GTGTTTGAGTACACGAAGGACGAGCTCTCAAAGGCAGTCGAGGACGCTGACCTTGTGGTG   240
Tx2.MDH      GTGACTGAGTACACGAAGGACGAGCTCTCAAAGGCAGTCGAGGGCGTTGACCTTGTGGTG   240
Tx3.MDH      GTGTTTGAGTACACGAAGGACGAGCTCTCAAAGGCAGTCGAGGACGCTGACCTTGTGGTG   240
             ***  ************************************** ** *************

Ref.MDH      ATTCCCGCTGGCGTGCCCCGTAAGCCGGGGATGACGCGCGACGACCTCTTCAACACGAAT   300
Tx2.MDH      ATTCCCGCTGGCGTGCCCCGTAAGCCGGGGATGACGCGCGACGACCTCTTCCACACGAAC   300
Tx3.MDH      ATTCCCGCTGGCGTGCCCCGTAAGCCGGGGATGACGCGCGACGACCTCTTCCACACGAAT   300
             *************************************************** ******* 

Ref.MDH      GCCAGCATCGTGCGCGATCTCTCAAAGGCTGTCGGCAAGGCCTCACCGAAGGCTATCATC   360
Tx2.MDH      GCCAGCATCGTGCGCGATCTCTCAAAGGCTGCCGGCAAGGCCTCACCGAAGGCTATCATC   360
Tx3.MDH      GCCAGCATCGTGCGCGATCTCTCAAAGGCTGTCGGCAAGGCCTCACCGAAGGCTATCATC   360
             ******************************* ****************************

Ref.MDH      GGTATCATCAGCAACCCTGTCAACAGCACTGTGCCCGTGGCTGCTGAGGCGCTGAAGGAG   420
Tx2.MDH      GGTATCATCAGCAACCCGGTCAACAGCACTGTGCCCGTGGCTGCTGAGGCGCTGAAGGAG   420
Tx3.MDH      GGTATCATCAGCAACCCTGTCAACAGCACTGTGCCCGTGGCTGCTGAGGCGCTGAAGGAG   420
             ***************** ******************************************

Ref.MDH      TTCGCGTACGATCCTGCGCGCCTCTTTGGCGTTACCACACTCGACGCTGTCCGTGCCCGC   480
Tx2.MDH      TTCGCGTACGATCCTGCGCGCCTCTTTGGCGTTACCACACTCGACGCTGTCCGTGCCCGC   480
Tx3.MDH      TTCGCGTACGATCCTGCGCGCCTCTTTGGCGTTACCACACTCGACGCTGTCCGTGCCCGC   480
             ************************************************************

Ref.MDH      ACCTTCGTCGCAGAGGCGCTCGGCGCAAGCCCGTACGACGTTAACGTCCCTGTTATTGGC   540
Tx2.MDH      ACCTTCGTCGCAGAGGCGCTCGGCGCAAGCCCGTGCGACGTTGACGTCCCTGTTATTGGC   540
Tx3.MDH      ACCTTCGTCGCAGAGGCGCTCGGCGCAAGCCCGTACGACGTTGACGTCCCTGTTATTGGC   540
             ********************************** ******* *****************

Ref.MDH      GGGCATAGCGGTGAGACGATTGTGCCGCTGCTCTCGGGCTTCCCGTCGCTGTCGGAGGAC   600
Tx2.MDH      GGGCATAGCGGTGAGACGATTGTGCCGCTGCTCTCGGGCTTCCCGTCGCTGTCGGAGGAC   600
Tx3.MDH      GGGCATAGCGGTGAGACGATTGTGCCGCTGCTCTCGGGCTTCCCGTCGCTGTCGGAGGAC   600
             ************************************************************

Ref.MDH      CAGGTGCGGCAGCTGACGCACCGCATTCAGTTCGGTGGTGACGAGGTGGTGAAGGCCAAG   660
Tx2.MDH      CAGGTGCGGCAGCTGACGCACCGCATTCAGTTCGGTGGTGACGAGGTGGTGAAGGCCAAG   660
Tx3.MDH      CAGGTGCGGCAGCTGACGCACCGCATTCAGTTCGGTGGTGACGAGGTGGTGAAGGCCAAG   660
             ************************************************************
```

```
Ref.MDH      GAAGGTGCGGGCTCGGCGACGCTGTCCATGGCGTACGCGGGCAACGAGTGGACGACGGCG    720
Tx2.MDH      GAAGGTGCGGGCTCGGCGACGCTGTCCATGGCGTACGCGGGCAACGAGTGGACGACGGCG    720
Tx3.MDH      GAAGGTGCGGGCTCGGCGACGCTGTCCATGGCGTACGCGGGCAACGAGTGGACGACGGCG    720
             ************************************************************

Ref.MDH      ATACTGCGCGCCCTCAATGGCGAGAAGGGTGTCGTGGTCTGCACGTACGTGCAGAGCTGC    780
Tx2.MDH      ATACTGCGCGCCCTCAATGGCGAGAAGGGTGTCGTGGTCTGCACGTACGTGCAGAGCTGC    780
Tx3.MDH      ATACTGCGCGCCCTCAATGGCGAGAAGGGTGTCGTGGTCTGCACGTACGTGCAGAGCTGC    780
             ************************************************************

Ref.MDH      GTGGAGCCGTCGTGCGCCTTCTTCAGCTCGCCGGTATTGCTGGGCAAGCGGGGTGTCGAA    840
Tx2.MDH      GTGGAGCCGTCGTGCGCCTTCTTCAGCTCGCCGGTATTGCTGGGCAAGCGGGGTGTCGAA    840
Tx3.MDH      GTGGAGCCGTCGTGCGCCTTCTTCAGCTCGCCGGTATTGCTGGGCAAGCGGGGTGTCGAA    840
             ************************************************************

Ref.MDH      AAGATCTACCCTGTGCCGACGCTGAACATATACGAAGAGAAGCTGATGTCCAAGTGCTTG    900
Tx2.MDH      AAGATTTACCCTGTGCCGACGCTGAACGCATACGAAGAGAAGCTGATGTCCAAGTGCTTG    900
Tx3.MDH      AAGATCTACCCTGTGCCGACGCTGAACATATACGAAGAGAAGCTGATGTCCAAGTGCTTG    900
             ***** *********************  *******************************

Ref.MDH      AAGGTTCTGCCGGGCGACATCAAGAAGGGCATCGAGCTCGGCAACTGGTAA     951
Tx2.MDH      AAGGTTCTGCCGGGCGACATCTAGAAGGGCATCGAGCTCGGCAACAGGTAA     951
Tx3.MDH      AAGGTTCTGCCGGGCGACATCAAGAAGGGCATCGAGCTCGGCAACTGGTAA     951
             ********************* *********************** *****
```

**MPI** CLUSTAL O(1.2.4) multiple sequence alignment

```
Ref.MPI      ATGTCTGAGCTCGTCAAGCTTGACGTGGGCCACCAGGACTATGCCTGGGGCAAGGATGCC   60
Tx2.CDS      ATGTCTGAGCTCGTCAAGCTTGACGTGGGCCACCAGGACTATGCCTGGGGCAAGGATGCC   60
Tx3.MPI      ATGTCTGAGCTCGTCAAGCTTGACGTGGGCCACCAGGACTATGCCTGGGGCAAGGATGCC   60
             ************************************************************

Ref.MPI      GCGTCCAGCTTCGTGGCGAAGATGAAGGGGTTGACGAACGACAAGTCCGGTAAGATGTTT   120
Tx2.CDS      GCGTCCAGCTTCGTGGCGAAGATGAAGGGGTTGACGAACGACAAGTCCGGTAAGATGTTT   120
Tx3.MPI      GCGTCCAGCTTCGTGGCGAAGATGAAGGGGTTGACGAACGACAAGTCCGGTAAGATGTTT   120
             ************************************************************

Ref.MPI      GCCGAGTTGTGGGTAGGCACGCACCTCAACTGCCCGTCGAAGATCGCCGACGGCAACGCG   180
Tx2.CDS      GCCGAGTTGTGGGTAGGCACGCACCTCAACTGCCCGTCGAAGATCGCCGACGGCAACGCG   180
Tx3.MPI      GCCGAGTTGTGGGTAGGCACGCACCTCAACTGCCCGTCGAAGATCGCCGACGGCAACGCG   180
             ************************************************************

Ref.MPI      CAGCTGCTCGAGGACTTCCTGAAGCAGCCAGAGAATAAGAAGAAGTACTTTTCGGAAGCT   240
Tx2.CDS      CAGCTGCTCGAGGACTTCCTGAAGCAGCCAGAGAATAAGAAGAAGTACTTTTCGGAAGCT   240
Tx3.MPI      CAGCTGCTCGAGGACTTCCTGAAGCAGCCAGAGAATAAGAAGAAGTACTTTTCGGAAGCT   240
             ************************************************************

Ref.MPI      CACCAGGCTACGACCTTCCGTGACACTGTGCCGTACCTGCTGAAGATTCTGTCGATTCGC   300
Tx2.CDS      CACCAGGCTACAACCTTCCGTGACACTGTGCCGTACCTGCTGAAGATTCTGTCGATTCGC   300
Tx3.MPI      CACCAGGCTACGACCTTCCGTGACACTGTGCCGTACCTGCTGAAGATTCTGTCGATTCGC   300
             *********** ************************************************

Ref.MPI      ACGGCTCTGTCGATTCAGGCGCACCCGTGCAAGAAGTTTGCCGAGGAGCTGCACGCTGCG   360
Tx2.CDS      ACGGCTCTGTCGATTCAGGCGCACCCGTGCAAGAAGTTTGCCGAGGAGCTGCACGCTGCG   360
Tx3.MPI      ACGGCTCTGTCGATTCAGGCGCACCCGTGCAAGAAGTTTGCCGAGGAGCTGCACGCTGCG   360
             ************************************************************

Ref.MPI      AGGCCGGATAAGTACAAGGACCCGAACCACAAGCCTGAGCTCATTTGCGCCTTGACCCCC   420
Tx2.CDS      AGGCCGGATAAGTACAAGGACCCGAACCACAAGCCTGAGCTCATTTGCGCCTTGACTCCC   420
Tx3.MPI      AGGCCGGATAAGTACAAGGACCCGAACCACAAGCCTGAGCTCATTTGCGCCTTGACCCCC   420
             *************************************** **************** ***

Ref.MPI      TTTGAGGCACTCTGCTGCTTCCGACCGCTCGGGGCCATCATCGCGTATCTGAAGCGCATT   480
Tx2.CDS      TTTGAGGCACTCTGCTGCTTCCGACCGCTCGGGGCCATCATCGCGTATCTGAAGCGCATT   480
Tx3.MPI      TTTGAGGCACTCTGCTGCTTCCGACCGCTCGGGGCCATCATCGCGTATCTGAAGCGCATT   480
             ************************************************************

Ref.MPI      CCAGAGCTGGCTGAGCTTGTAGGCGCCGAAGCGGTGCTGGGTCAGTACATGATGGCGCCG   540
Tx2.CDS      CCTGAGCTGGCAGAGCTTGTGGGTGCCGAAGCGGTGCTGGGTCAGTACGTGATGGCGCCG   540
Tx3.MPI      CCAGAGCTGGCTGAGCTTGTAGGCGCCGAAGCGGTGCTGGGTCAGTACATGATGGCGCCG   540
             ** ******** ******** ** ************************ ***********

Ref.MPI      GAGACCGCGCTGCCTGCAACCGACAGCGACGAGGAGAAGCAGTTACTGAAGACTATGATG   600
Tx2.CDS      GAGAGCGCGCTGCCTGCAACCGACAGCGACGAGGAGAAGCAGTTACTGAAGGCTATGATG   600
Tx3.MPI      GAGACCGCGCTGCCTGCAACCGACAGCGACGAGGAGAAGCAGTTACTGAAGGCTATGATG   600
             **** ********************************************** ********

Ref.MPI      ACGACCGTGTATGCGGCCGCGGATGACGTCGTCACGAAGGCGCTGCGTCTGCACCTTCAG   660
Tx2.CDS      ACGAACGTGTACGCGGCCGCGGATGACGTCGTCACGAAGGCGCTGCGTCTGCACCTTCAG   660
Tx3.MPI      ACGACCGTGTACGCGGCCGCGGATGACGTCGTCACGAAGGCGCTGCGTCTGCACCTTCAG   660
             **** ****** ************************************************

Ref.MPI      TGCATCGAGGAGAAGGGTGCGCAGTGCGCCGAAGACGAGCTCTTTGCTCGTATTTACAGG   720
Tx2.CDS      CGGATCGAGGAGAAGGGTGCGCAGTGCGCCGAGGACGAGCTCTTTGCTCGCATTTACAGA   720
Tx3.MPI      CGCATCGAGGAGAAGGGTGCGCAGTGCGCCGAAGACGAGCTCTTTGCTCGTATTTACAGG   720
              * ***************************** ***************** ******** 
```

```
Ref.MPI     CAGTACCCGGATGATGTCGGGTGCTGGATGGTTTACTTCCTCAATTACGTACAGATGGTG   780
Tx2.CDS     CAGTACCCGGATGATGTCGGGTGCTGGATGGTTTACTTCCTCAATTACGTACAGATGGTG   780
Tx3.MPI     CAGTACCCGGATGATGTCGGGTGCTGGATGGTTTACTTCCTCAATTACGTACAGATGGTG   780
            ************************************************************

Ref.MPI     CCCGGGGAGGCTCTCTTCCTATCGGACAGCGAGCCGCACGCATATATCAGCGGCGACGGT   840
Tx2.CDS     CCCGGGGAGGCTCTCTTCCTATCGGACAGCGAGCCGCACGCATATATCAGCGGCGACGGT   840
Tx3.MPI     CCCGGGGAGGCTCTCTTCCTATCGGACAGCGAGCCGCACGCATATATCAGCGGCGACGGT   840
            ************************************************************

Ref.MPI     GTCGAGATCATGGCATGCAGCGACAACGTCGTGCGTGCTGGACTCACGCCGAAGTGGAAG   900
Tx2.CDS     GTCGAGATCATGGCATGCAGCGACAACGTAGTGCGTGCTGGACTCACGCCGAAGTGGAAG   900
Tx3.MPI     GTCGAGATCATGGCATGCAGCGACAACGTCGTGCGTGCTGGACTCACGCCGAAGTGGAAG   900
            ***************************** ******************************

Ref.MPI     GATGTGCCGACGCTCATAAGCATGTTGAAGTACGACACGACCGGGCTTGCGTCTGCCCGC   960
Tx2.CDS     GATGTGCCGACGCTCATAAGCATGTTGAAGTACGACACGACCGGGCTTGCGTCTGCCCGC   960
Tx3.MPI     GATGTGCCGACGCTCATAAGCATGTTGAAGTACGACACGACCGGGCTTGCGTCTGCCCGC   960
            ************************************************************

Ref.MPI     CACGAGAAGAAGATCGGCGAGGACGCGGCGCAGTGGCAGGTGCAGTATTACCAACCACCG   1020
Tx2.CDS     CACGAGAAGAAGATCGGCGAGGACGCGGCGCAGTGGCAGGTGCAGTATTACCAACCACCG   1020
Tx3.MPI     CACGAGAAGAAGATCGGCGAGGACGCGGCGCAGTGGCAGGTGCAGTATTACCAACCACCG   1020
            ************************************************************

Ref.MPI     GCACAGTTCCCCGACTTTTCGCTGTACCGCCTGCAGTACGAGCATGCTTCAGGCAACGGG   1080
Tx2.CDS     GCACAGTTCCCGGACTTTTCGCTGTACCGCCTGCAGTACGAGCATGCTTCAGGCAACGGG   1080
Tx3.MPI     GCACAGTTCCCCGACTTTTCGCTGTACCGCCTGCAGTACGAGCATGCTTCAGGCAACGGG   1080
            *********** ************************************************

Ref.MPI     ATGACCTCCGTGACCCTGCCGACGATAGGCCTGGGTTTCTGCTTGGAGGGGTCTGCCAAG   1140
Tx2.CDS     ATGACCTCCGTGACCCTGCCGACGATAGGCCTGGGTTTCTGCTTGGAGGGGTCTGCCAAG   1140
Tx3.MPI     ATGACCTCCGTGACCCTGCCGACGATAGGCCTGGGTTTCTGCTTGGAGGGGTCTGCCAAG   1140
            ************************************************************

Ref.MPI     GTGAACGATACGACGGTGAACGCCGGCGACTGCTTTGCGGTGCCGTACGGCAAGCTCACG   1200
Tx2.CDS     GTGAACGGTACGACGGTGAACGCCGGCGACTGCTTTGCGGTACCGTACGGCAAGCTCACG   1200
Tx3.MPI     GTGAACGATACGACGGTGAACGCCGGCGACTGCTTTGCGGTGCCGTACGGCAAGCTCACG   1200
            ******* ********************************* ******************

Ref.MPI     TGCCAAGCCGAGGGAGCGAAGGCGCTTGTGTTTGTTGCGTCGACCAACGACTTGAGCGAC   1260
Tx2.CDS     TGCCAAGCCGAGGGAGCGAAGGCGCTTGTGTTTGTTGCGTCGACCAACGACTTGAGCGAC   1260
Tx3.MPI     TGCCAAGCCGAGGGAGCGAAGGCGCTTGTGTTTGTTGCGTCGACCAACGACTTGAGCGAC   1260
            ************************************************************

Ref.MPI     AAGTAA      1266
Tx2.CDS     AAGTAA      1266
Tx3.MPI     AAGTAA      1266
            ******
```

**PGD** CLUSTAL O(1.2.4) multiple sequence alignment

```
PGD.ref      ATGTCGAACGACCTCGGTATTATCGGTCTCGGCGTCATGGGCGCGAATCTCGCCCTGAAC   60
Tx2.PGD      ATGTCGAACGACCTCGGTATTATCGGTCTCGGCGTCATGGGCGCGAATCTCGCCCTGAAC   60
Tx3.PGD      ATGTCGAACGACCTCGGTATTATCGGTCTCGGCGTCATGGGCGCGAATCTCGCCCTGAAC   60
             ************************************************************

PGD.ref      ATCGCCGAGAAGGGATTTAAAGTTGCCGTCTTCAACCGCACCTACACGAAGACGACGTCG   120
Tx2.PGD      ATCGCCGAGAAGGGATTTAAAGTTGCCGTCTTCAACCGCACCTACACGAAGACGACGTCG   120
Tx3.PGD      ATCGCCGAGAAGGGATTTAAAGTTGCCGTCTTCAACCGCACCTACACGAAGACGACGTCG   120
             ************************************************************

PGD.ref      TTTCTCAAGGAGCATGAGAACGAGGCGCTTGCCGTCAACCTAAAGGGGTACGAGACAATG   180
Tx2.PGD      TTTCTCAAGGAGCATGAGAACGAGGCGCTTGCCGTCAACCTAAAGGGGTACGAGACAATG   180
Tx3.PGD      TTTCTCAAGGAGCATGAGAACGAGGCGCTTGCCGTCAACCTAAAGGGGTACGAGACAATG   180
             ************************************************************

PGD.ref      AAGGAGTTCGCCGCGTCCCTCAAGAAGCCGCGCCGCGCGTTCATTCTCGTCCAGGCCGGC   240
Tx2.PGD      AAGGAGTTCGCCGCGTCCCTCAAGAAGCCGCGCCGCGCGTTCATTCTCGTCCAGGCCGGC   240
Tx3.PGD      AAGGAGTTCGCCGCGTCCCTCAAGAAGCCGCGCCGCGCGTTCATTCTCGTCCAGGCCGGC   240
             ************************************************************

PGD.ref      GCTGCTACGGACTCCACGATCGAACAGCTCAAGGAAGTTTTCGAGGAGGGCGACATCGTC   300
Tx2.PGD      GCTGCTACGGACTCCACGATCGAACAGCTCAAGGAAGTTTTCGAGGAGGGCGACATCGTC   300
Tx3.PGD      GCTGCTACGGACTCCACGATCGAACAGCTCAAGGAAGTTTTCGAGGAGGGCGACATCGTC   300
             ************************************************************

PGD.ref      ATAGACACTGGCAATGCGAACTTCAAGGACCAGGACAGGCGCGCCGCGCAGTTGGAGAGC   360
Tx2.PGD      ATAGACACTGGCAATGCGAACTTCAAGGACCAGGACAGGCGCGCCGCGCAGTTGGAGAGC   360
Tx3.PGD      ATAGACACTGGCAATGCGAACTTCAAGGACCAGGACAGGCGCGCCGCGCAGTTGGAGAGC   360
             ************************************************************

PGD.ref      CAGGGTCTCCGCTTCCTCGGCATGGGCATCTCCGGTGGTGAGGAGGGTGCGCGCAAGGGG   420
Tx2.PGD      CAGGGTCTCCGCTTCCTCGGCATGGGCATCTCCGGTGGTGAGGAGGGTGCGCGCAAGGGG   420
Tx3.PGD      CAGGGTCTCCGCTTCCTCGGCATGGGCATCTCCGGTGGTGAGGAGGGTGCGCGCAAGGGG   420
             ************************************************************

PGD.ref      CCGGCCTTCTTCCCTGGGGGCACACCAAGCGTGTGGGAGGAGGTACGCGCGATCGTGGAG   480
Tx2.PGD      CCGGCCTTCTTCCCTGGGGGCACACCAAGCGTGTGGGAGGAGGTACGCGCGATCGTGGAG   480
Tx3.PGD      CCGGCCTTCTTCCCTGGGGGCACACCAAGCGTGTGGGAGGAGGTACGCGCGATCGTGGAG   480
             ************************************************************

PGD.ref      GCGGCTGCGGCCAAGGCTGAGGACGGTCGCCCGTGCGTGACGTTCAACGGCAAGGGCGGC   540
Tx2.PGD      GCGGCTGCGGCCAAGGCTGAGGACGGTCGCCCGTGCGTGACGTTCAACGGCAAGGGCGGC   540
Tx3.PGD      GCGGCTGCGGCCAAGGCTGAGGACGGTCGCCCGTGCGTGACGTTCAACGGCAAGGGCGGC   540
             ************************************************************


PGD.ref      GCTGGATCCTGCGTGAAGATGTACCACAACGCTGGTGAGTACGCCGTGCTGCAGATCTGG   600
Tx2.PGD      GCTGGATCCTGCGTGAAGATGTACCACAACGCTGGTGAGTACGCCGTGCTGCAGATCTGG   600
Tx3.PGD      GCTGGATCCTGCGTGAAGATGTACCACAACGCTGGTGAGTACGCCGTGCTGCAGATCTGG   600
             ************************************************************

PGD.ref      GGTGAGGCGTACAGCGCCTTGCTTGCCTTCGGATTCAACAACGATCAGATCGCTGACGTG   660
Tx2.PGD      GGTGAGGCGTACAGCGCCTTGCTTGCCTTCGGATTCAACAACGATCAGATCGCTGACGTG   660
Tx3.PGD      GGTGAGGCGTACAGCGCCTTGCTTGCCTTCGGATTCAACAACGATCAGATCGCTGACGTG   660
             ************************************************************

PGD.ref      TTCGAGTCGTGGAAGGCAGACGGCTTCCTCAAATCCTACATGCTCGACATCTCTATTGTC   720
Tx2.PGD      TTCGAGTCGTGGAAGGCAGACGGCTTCCTCAAATCCTACATGCTCGACATCTCTATTGTC   720
Tx3.PGD      TTCGAGTCGTGGAAGGCAGACGGCTTCCTCAAATCCTACATGCTCGACATCTCTATTGTC   720
             ************************************************************
```

```
PGD.ref      GCTTGCCGGGCGAGGGAGGCAGCAGGCAACTATCTGTCAGAGAAGGTGTTGGACCGCATC   780
Tx2.PGD      GCTTGCCGTGCGAGGGAGGCAGCAGGCAACTATCTGTCAGAGAAGGTGTTGGACCGCATC   780
Tx3.PGD      GCTTGCCGGGCGAGGGAGGCAGCAGGCAACTATCTGTCAGAGAAGGTGTTGGACCGCATC   780
             ******** ***************************************************

PGD.ref      GGCTCCAAAGGTACCGGCCTGTGGTCGGCCCAGGAGGCTCTGGAAGTTGGGGTGCCAGCG   840
Tx2.PGD      GGCTCCAAAGGTACCGGCCTGTGGTCGGCCCAGGAGGCTCTGGAAGTTGGGGTGCCAGCG   840
Tx3.PGD      GGCTCCAAAGGTACCGGCCTGTGGTCGGCCCAGGAGGCTCTGGAAGTTGGGGTGCCAGCG   840
             ************************************************************

PGD.ref      CCCTCGCTCAACATGGCCGTCATCTCACGCCAGATGACCATGTACAAAGCGGAGCGTGTC   900
Tx2.PGD      CCCTCGCTCAACATGGCCGTCATCTCACGCCAGATGACCATGTACAAAGCGGAGCGTGTC   900
Tx3.PGD      CCCTCGCTCAACATGGCCGTCATCTCACGCCAGATGACCATGTACAAAGCGGAGCGTGTC   900
             ************************************************************

PGD.ref      GCGAACTCCAAGGCATTCCCTCACTTCCCTTGCGGCCCGTGTGAGAAAGCCACGGACAAG   960
Tx2.PGD      GCGAACTCCAAGGCATTCCCTCACTTCCCTTGCGGCCCGTGTGAGAAAGCCACGGACAAG   960
Tx3.PGD      GCGAACTCCAAGGCATTCCCTCACTTCCCTTGCGGCCCGTGTGAGAAAGCCACGGACAAG   960
             ************************************************************

PGD.ref      TCCCCGAACTCCCCCGAAGCGAAGCAGCTATTCCACGCCGTCAGCCTTTCCATCATTGCA   1020
Tx2.PGD      TCCCCGAACTCCCCCGAAGCGAAGCAGCTGTTCCACGCCGTCAGCCTTTCCATCATTGCA   1020
Tx3.PGD      TCCCCGAACTCCCCCGAAGCGAAGCAGCTGTTCCACGCCGTCAGCCTTTCCATCATTGCA   1020
             ***************************** ******************************

PGD.ref      AGTTACGCGCAGATGTTCCAGTGCTTGCGCGAGCTGGACAAGGTGTACGGATTTGGCTTG   1080
Tx2.PGD      AGTTACGCGCAGATGTTCCAGTGCTTGCGCGAGCTGGACAAGGTGTACGGATTTGGCTTG   1080
Tx3.PGD      AGTTACGCGCAGATGTTCCAGTGCTTGCGCGAGCTGGACAAGGTGTACGGATTTGGCTTG   1080
             ************************************************************

PGD.ref      AACCTGCCCGCCACCATCGCAACCTTCCGCGCCGGCTGCATTCTGCAGGGCTACCTGCTG   1140
Tx2.PGD      AACCTGCCCGCCACCATCGCAACCTTCCGCGCCGGCTGCATTCTGCAGGGCTACCTGCTG   1140
Tx3.PGD      AACCTGCCCGCCACCATCGCAACCTTCCGCGCCGGCTGCATTCTGCAGGGCTACCTGCTG   1140
             ************************************************************

PGD.ref      GGGCCTATGACGAAGGCCTTCGAGGAGAACCCGAACCTGCCCAACCTGTTGGACGCCTTC   1200
Tx2.PGD      GGGCCTATGACGAAGGCCTTCGAGGAGAACCCGAACCTGCCCAACCTGTTGGACGCCTTC   1200
Tx3.PGD      GGGCCTATGACGAAGGCCTTCGAGGAGAACCCGAACCTGCCCAACCTGTTGGACGCCTTC   1200
             ************************************************************

PGD.ref      ACCAAGGAGATAGCTGCGGGCCTTAATGACTGCCGCCAGATTCTCGCCAGGCTCACAGTG   1260
Tx2.PGD      ACCAAGGAGATAGCTGCGGGCCTTAATGACTGCCGCCAGATTCTCGCCAGGCTCACAGTG   1260
Tx3.PGD      ACCAAGGAGATAGCTGCGGGCCTTAATGACTGCCGCCAGATTCTCGCCAGGCTCACAGTG   1260
             ************************************************************

PGD.ref      AATACGGCAGTGTCACTGCCGGTCATGATGGCCTCACTCTCCTACATCAACGCCATGTAC   1320
Tx2.PGD      AATACGGCAGTGTCACTGCCGGTCATGATGGCCTCACTCTCCTACATCAACGCCATGTAC   1320
Tx3.PGD      AATACGGCAGTGTCACTGCCGGTCATGATGGCCTCACTCTCCTACATCAACGCCATGTAC   1320
             ************************************************************

PGD.ref      ACGGAGATTCTTCCGTACGGACAGCTGGTGTCGTTGCAGCGTGACGTCTTTGGCCGCCAC   1380
Tx2.PGD      ACGGAGATTCTTCCGTACGGACAGCTGGTGTCGTTGCAGCGTGACGTCTTTGGCCGCCAC   1380
Tx3.PGD      ACGGAGATTCTTCCGTACGGACAGCTGGTGTCGTTGCAGCGTGACGTCTTTGGCCGCCAC   1380
             ************************************************************

PGD.ref      GGCTACGAGCGCACAGACAGGGATGGTCGTGAGTCGTTTGAATGGCCCGCACTGCAGTAG   1440
Tx2.PGD      GGCTACGAGCGCACAGACAGGGATGGTCGTGAGTCGTTTGAATGGCCCGCACTGCAGTAG   1440
Tx3.PGD      GGCTACGAGCGCACAGACAGGGATGGTCGTGAGTCGTTTGAATGGCCCGCACTGCAGTAG   1440
             ************************************************************
```
